# Effect of clustering and correlation of belief systems on infectious disease outbreaks

**DOI:** 10.1101/2020.12.08.20246298

**Authors:** Claus Kadelka, Audrey McCombs

## Abstract

Contact between people with similar opinions and characteristics occurs at a higher rate than among other people, a phenomenon known as homophily. The presence of clusters of unvaccinated people has been associated with increased incidence of infectious disease outbreaks despite high vaccination coverage. The epidemiological consequences of homophily regarding other beliefs and correlations among belief systems are however poorly understood. Here, we use a simple compartmental disease model as well as a more complex COVID-19 model to study how homophily and correlation of belief systems in a social interaction network affect the probability of disease outbreak and COVID-19-related mortality. We find that the current social context, characterized by the presence of homophily and correlations between who vaccinates, who engages in risk reduction, and individual risk status, corresponds to a situation with substantially worse disease burden than in the absence of heterogeneities. In the presence of an effective vaccine, relative effects of homophily and correlation of belief systems become stronger. Further, the optimal vaccination strategy depends on the degree of homophily regarding vaccination status as well as the relative level of risk mitigation high- and low-risk individuals practice. The developed methods are broadly applicable to any investigation in which node attributes in a graph might reasonably be expected to cluster or exhibit correlations.

## Introduction

Infectious disease outbreaks have been on the rise for several decades, and account for more than one in eight deaths globally [1]. A comprehensive study of an infectious disease outbreak such as the current COVID-19 pandemic must involve not only the biological properties of the disease and its causal pathogen but also the societal circumstances affecting disease spread. Classical differential equation models assume homogeneous mixing (i.e., random contacts) of individuals and fail to account for the occurrence of increased interactions among people with similar beliefs or circumstances, a phenomenon known as homophily. Network models, on the other hand, allow for the study of the effect of homophily on disease outbreaks by explicitly incorporating clustering of individuals with similar beliefs into social or physical interaction networks. Clustering of individuals who choose not to vaccinate for a variety of reasons (religious beliefs, fear of side effects, etc.) has been widely observed [2, 3, 4], and this type of homophily has been associated with both more frequent and larger disease outbreaks than would be expected under homogeneous or random social mixing [5, 6, 7]. Models that assume homogeneous mixing are therefore likely too optimistic and underestimate the level of vaccination required to achieve herd immunity and avoid outbreaks [8].

While the epidemiological implications of homophily regarding vaccination status have been well-studied, the effect of clustering of individuals with respect to other beliefs, such as trust in the effectiveness of social distancing measures, has not received as much attention [9]. Especially during a pandemic, when risk mitigation measures are implemented globally, homophily regarding various beliefs and correlations between them may have profound effects on disease spread. The correlation between social beliefs and partisan identification has been increasing in the United States [10], and ideological overlap between the two major political parties has diminished [11]. Recent polls by Gallup indicate that, in the United States, Democrats compared to Republicans are more likely to engage in risk mitigation (wearing masks [12], avoiding eating out [13], avoiding flying [14], etc.) and to receive a COVID-19 vaccine [15]. Given the increases in both opinion polarization and correlations among opinions, as well as the effect homophily can have on disease dynamics, further work on the epidemiological consequences of correlated and clustered belief systems is warranted.

Generating a network with *a priori* specified clustering and correlation among node attributes (belief in the safety of a COVID-19 vaccine, belief in the effectiveness of social distancing, etc.) can be technically challenging. In this study, we present a novel technique for applying binary attributes with a pre-defined correlation structure to a physical interaction network that exhibits a pre-defined level of homophily for each attribute. Using this technique, we investigate how homophily and correlations among several beliefs and circumstances affect the spread of an infectious disease in a Watts-Strogatz small-world physical interaction network (a community, a city, etc.) [16], and substantially influence the outcome of epidemiological studies and their predictions (Figure 1). First, we consider a simple compartmental infectious disease model with two binary belief attributes: confidence in vaccines and attitude toward social distancing measures. That is, a person either agrees to be vaccinated or not, and a person either engages in enhanced risk reduction (social distancing, mask wearing, etc.) or not. Then, in a more complex model developed specifically for COVID-19 [17], we add a third binary attribute distinguishing between high- and low-risk individuals.

**Figure 1:**
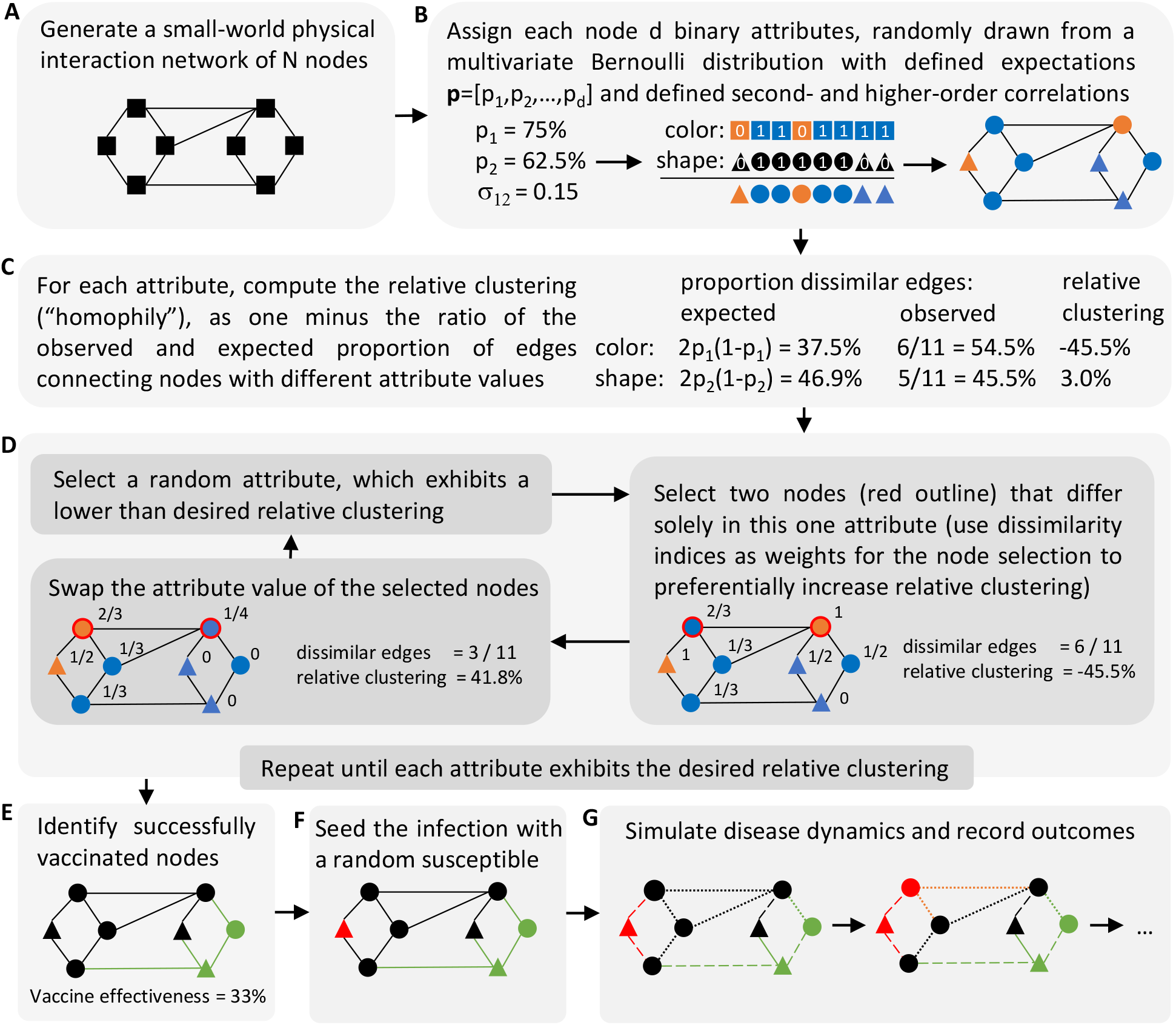
Graphical overview of a simulation run. (A) Generation of a physical interaction network, (B) Assignment of d correlated binary attribute values to each node. If d = 2, the attributes represent e.g. attitude towards vaccines (color) and social distancing (shape), Computation of the relative clustering level (a measure of homophily) of each attribute, Overview of the clustering algorithm used to assign attributes so that the network exhibits a desired level of homophily for each attribute. Values to the right of each node indicate its dissimilarity index: the proportion of neighbors with a different attribute value. (E) Vaccination of all nodes with a positive (blue) attitude towards vaccines and removal of those successfully vaccinated (green), (F) Infection of a randomly selected susceptible node (red), (G) Simulation of the spread of the infection and recording of outcomes. The likelihood of interaction (edge weight) depends on whether nodes practice social distancing (circles) or not (triangles).

## Results

The generic infectious disease model yielded several expected results. An increase in the proportion of vaccinated, an increase in the proportion of distancers, an increase in the vaccine effectiveness, and an increase in the level of distancing practiced all led to lower disease burden, quantified by the disease outbreak probability (Figure 2) as well as the initial basic reproductive number *R*_0_ (Figure S1). The two outcome measures were highly positively correlated; therefore, we focused primarily on the outbreak probability, which in network models incorporates more information than *R*_0_.

**Figure 2:**
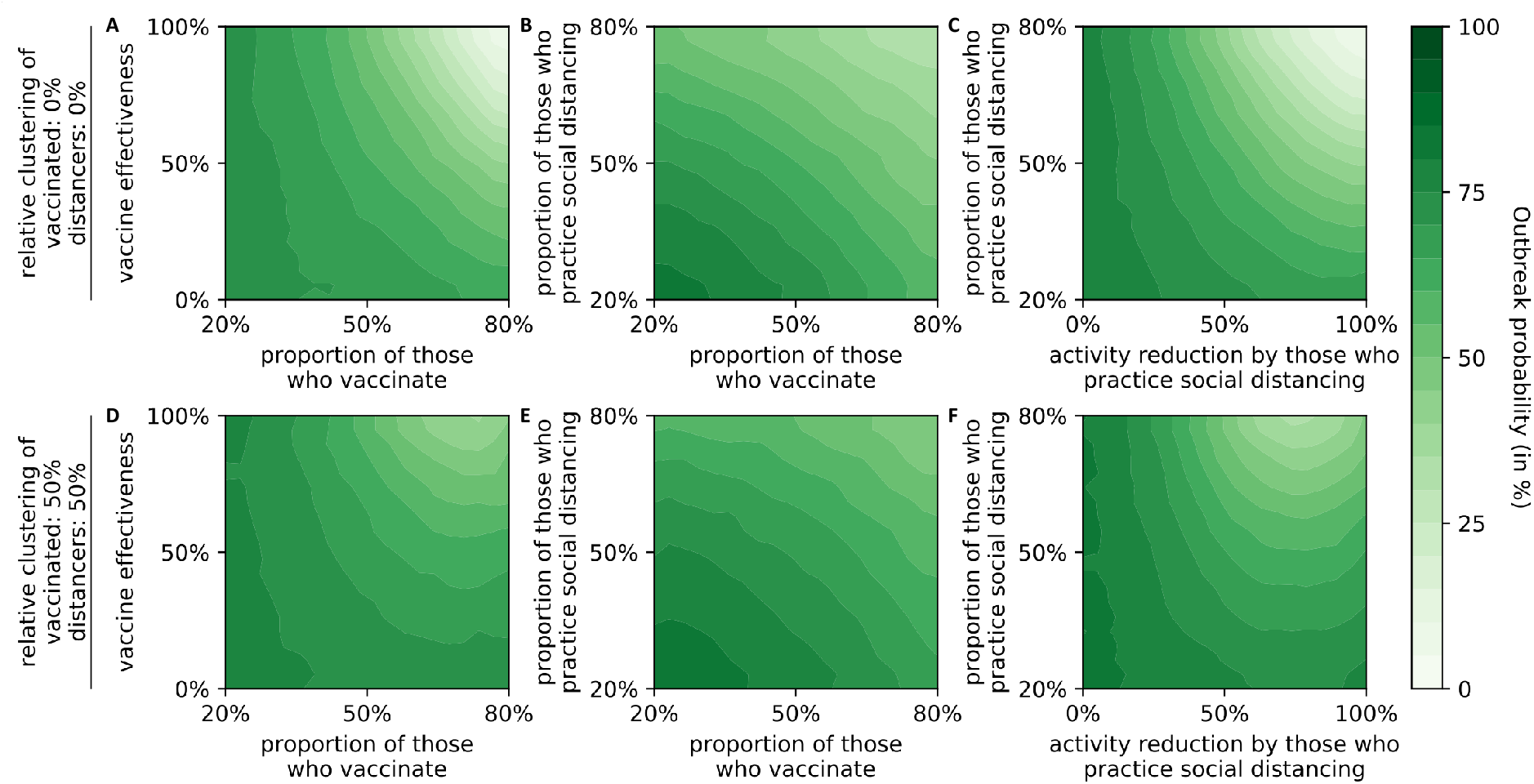
Effect of vaccine and social distancing parameters on outbreak probability. Contour plots were generated from 10,000,000 independent simulation runs with four vaccine and social distancing parameters chosen uniformly at random (axes show parameter ranges). Relative clustering levels of those who vaccinate and of those who practice distancing were fixed at (A-C) 0% and (D-F) 50%. Data was binned and smoothed using a two-dimensional Savitzky-Golay filter [18] (details in Methods).

Perfect isolation by those who practice social distancing led to more outbreaks than very high levels of distancing (Figure S1F). Similarly, in the case of a highly effective vaccine very high vaccine coverage (80%) led to more outbreaks than slightly lower coverage (Figure S1D). Both these counter-intuitive observations only occurred in the presence of clustering, and they are likely model artifacts: The activity level of each individual corresponds to the probability that this individual is chosen as the initially infected seed case. If the activity level of distancing individuals is positive (e.g. 75% reduction), then there remains a small chance that an individual who distances is chosen as the seed case. If this happens, an outbreak is highly unlikely if the distancers cluster together. At perfect isolation (100% reduction), solely non-distancers, who also cluster together in the presence of homophily, are chosen as seed cases. Similar reasoning explains the second observation.

Outbreaks occurred more frequently in interaction networks in which individuals with similar beliefs regarding vaccination and social distancing clustered (Figure S1D-F) than without clustering (Figure S1A-C). In order to obtain a detailed understanding of the effect of clustering and correlation of opinions on the outbreak probability, we fixed the values of several model parameters: we considered a situation where 2/3 of individuals vaccinate and 2/3 reduce their social contacts by 50%. As expected, outbreaks occurred less frequently in the presence of a more effective vaccine (Figure 3). Contact networks in which those who vaccinate were also more likely to socially distance (correlation= 0.45) exhibited more outbreaks, while a negative correlation (− 0.45) between vaccination and distancing led to fewer outbreaks than a situation with no correlation (Figure 3A,B). Contact networks in which those who vaccinate cluster were more likely to experience an outbreak, especially at higher levels of vaccine effectiveness (Figure 3C). Similarly, clustering of those who distance led to more outbreaks, but the effect was smaller (Figure 3D). Moreover, this effect became stronger as the correlation between vaccinated and distancers increased, and was absent when the correlation was negative. Interestingly, clustering of distancers led to slightly more outbreaks than no clustering in situations “without” a vaccine (effectiveness = 0%). In this case, as expected, it did not matter if those who received the vaccine clustered or not.

**Figure 3:**
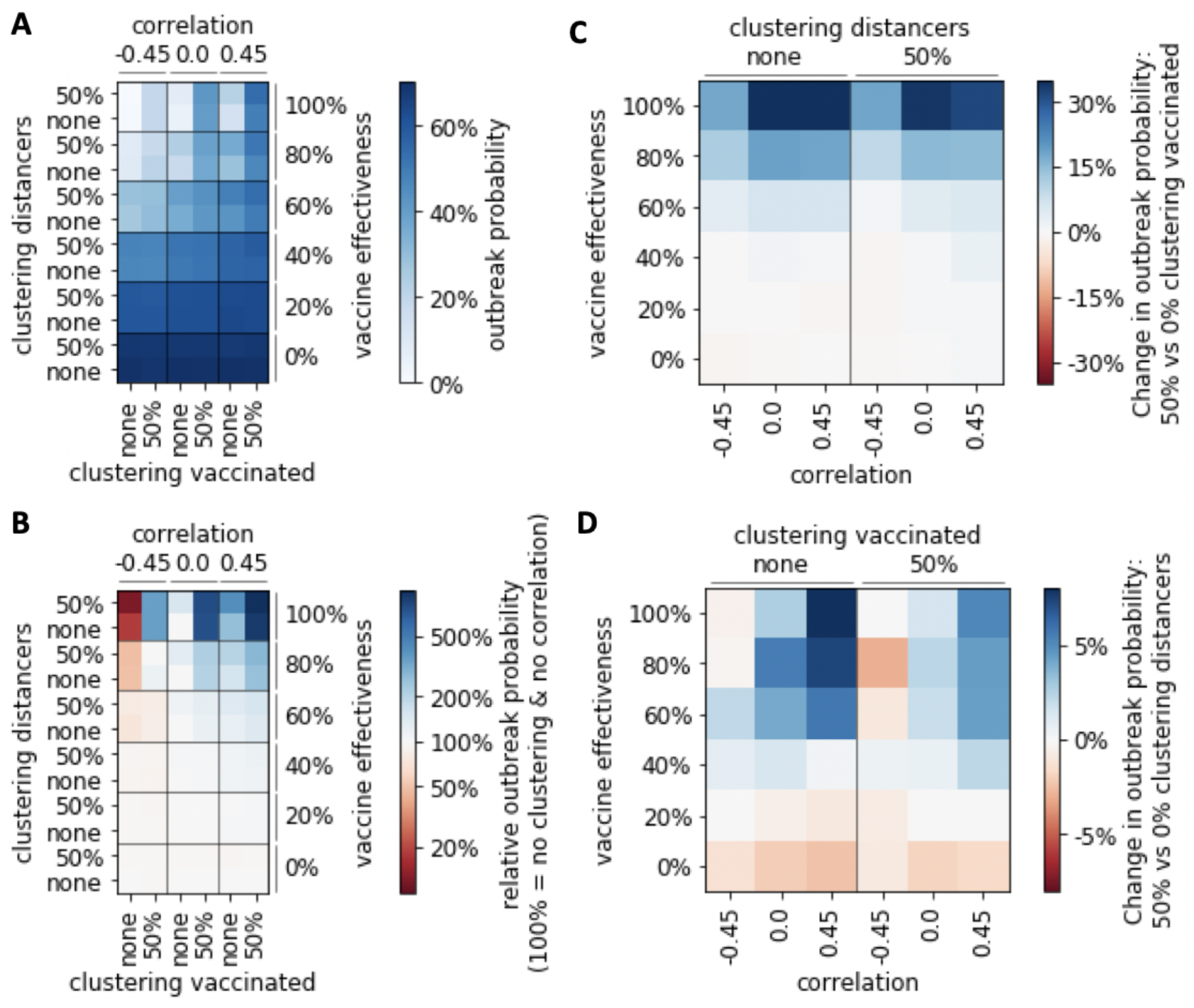
Effect of clustering and correlation of opinions on outbreak probability. (A-B) Outbreak probability under different scenarios with respect to clustering and correlation of those who vaccinate and those who distance, and for different levels of vaccine effectiveness. (A) absolute outbreak probability (in %), (B) relative change in outbreak probability compared to the homogeneous case of no clustering and no correlation. (C-D) Absolute change in outbreak probability when comparing physical interaction networks where (C) vaccinated, (D) distancers cluster versus networks without clustering of (C) vaccinated, (D) distancers.

After being vaccinated individuals may choose to increase their level of social contacts because they believe they are immune, a phenomenon known as risk compensation [19]. We considered a model scenario in which those who vaccinate increase their activity levels on average by up to 41%. The release of an ineffective vaccine coupled with increased activity levels led to more outbreaks than a situation without a vaccine (region to the left of the red line in Figure 4). In contact networks where those who vaccinate and those who distance clustered and coincided (correlation = 0.45; Figure 4A), vaccines needed to be slightly more effective to not be detrimental than in networks without such clustering and correlation (Figure 4B). These findings did not depend on the particular choice of the proportion of those who vaccinate and those who distance (Table S1).

**Figure 4:**
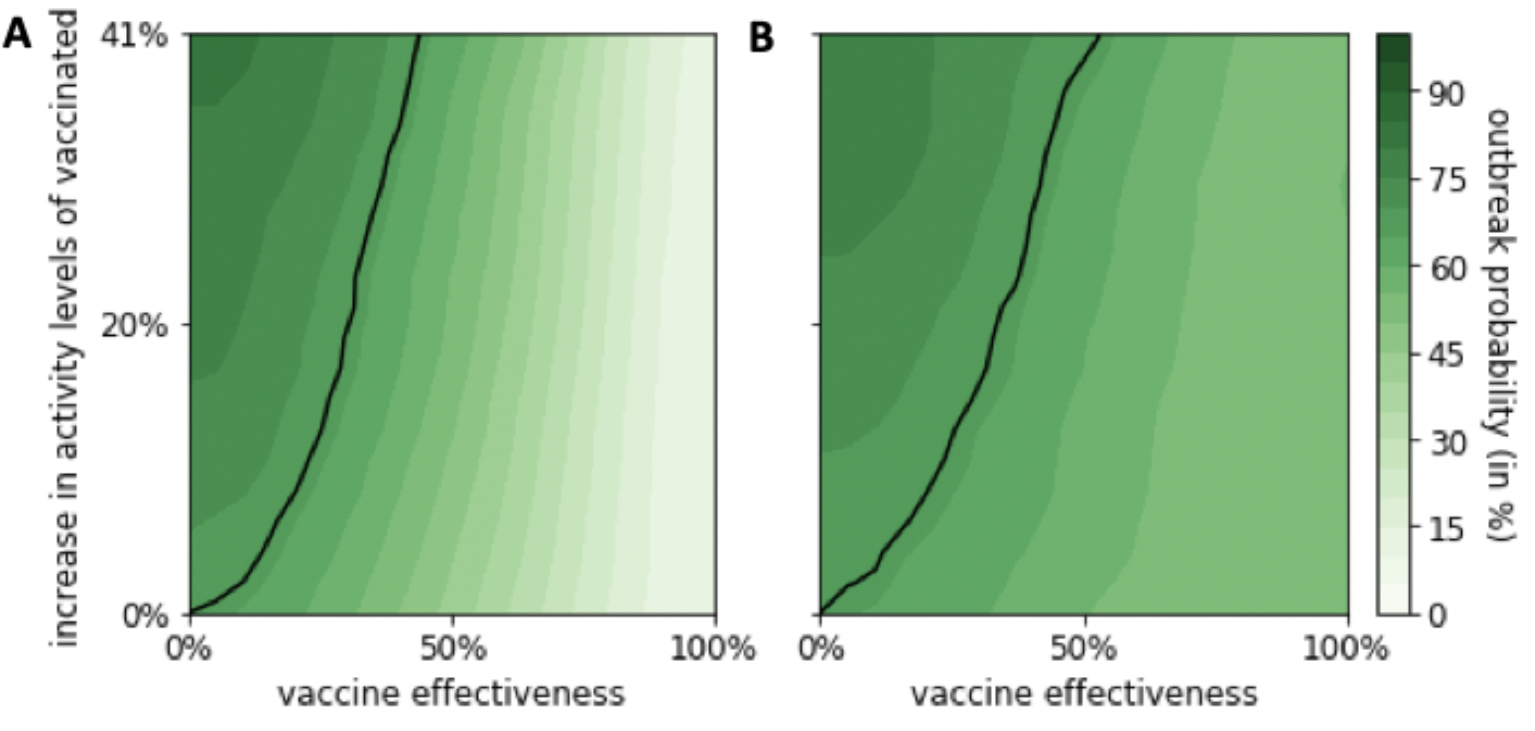
Effect of increased activity levels by individuals who have received a vaccine. The outbreak probability is shown for different levels of vaccine effectiveness (x-axis) and increased average activity levels by those who received a vaccine (y-axis). A black line depicts the x,y-coordinates at which the presence of the vaccine does not change the outbreak probability. To the left (right) of this line, the presence of the vaccine is detrimental (beneficial). Two different scenarios regarding clustering and correlation of those who vaccinate and those who distance are considered: (A) 0% relative clustering and no correlation, (B) 50% relative clustering of those who vaccinate and those who distance and 0.45 correlation. In both plots, a fixed proportion of 65% of all individuals receive a vaccine and 65% of all individuals practice social distancing, i.e., reduce their social contacts by 50%. Data was binned and smoothed using a two-dimensional Savitzky-Golay filter [18] (details in Methods). See Table S1 for a sensitivity analysis where these proportions are varied.

An important characteristic of COVID-19 is the increased risk of severe disease and death for older adults and people with comorbidities [20]. A recent COVID model captures this differential risk by distinguishing between low-risk (2/3 of all individuals in the United States) and high-risk individuals [17]. Here, we adapted this model to investigate the effects of both increased contact between individuals of the same risk group, as well as differential vaccination coverage and social distancing levels between risk groups. As the basic reproductive number and the outbreak probability both focus on disease transmission and fail to describe the differential risk of severe disease and death, for this analysis we focused instead on the number of deaths due to COVID-19.

In the case of a highly effective vaccine, clustering of those who vaccinate remained the most important variable: with a perfect vaccine (100% effective), almost 600% more people die when comparing 50% with 0% relative clustering of vaccinated individuals (Figure 5A,B). Clustering of distancers had only a small effect on mortality, and the direction of the effect was dependent on the vaccine effectiveness: In the presence of a bad vaccine (effectiveness ≤ 40%), slightly fewer people died when those who practiced social distancing clustered, while the opposite was true in situations with a more effective vaccine (effectiveness ≥ 60%). If those who vaccinated were also more likely to practice social distancing (correlation = 0.15), more deaths occurred, while mortality was reduced if individuals who did not get vaccinated were more likely to practice social distancing (correlation = − 0.15). Whether or not high-risk individuals clustered had very little effect on the number of deaths. However, the correlations between risk status, vaccination and social distancing proved important (Figure 5A-C). With increasing vaccine effectiveness, overall mortality decreased when high-risk individuals were more likely to get vaccinated and practice social distancing, although the latter effect was weaker.

**Figure 5:**
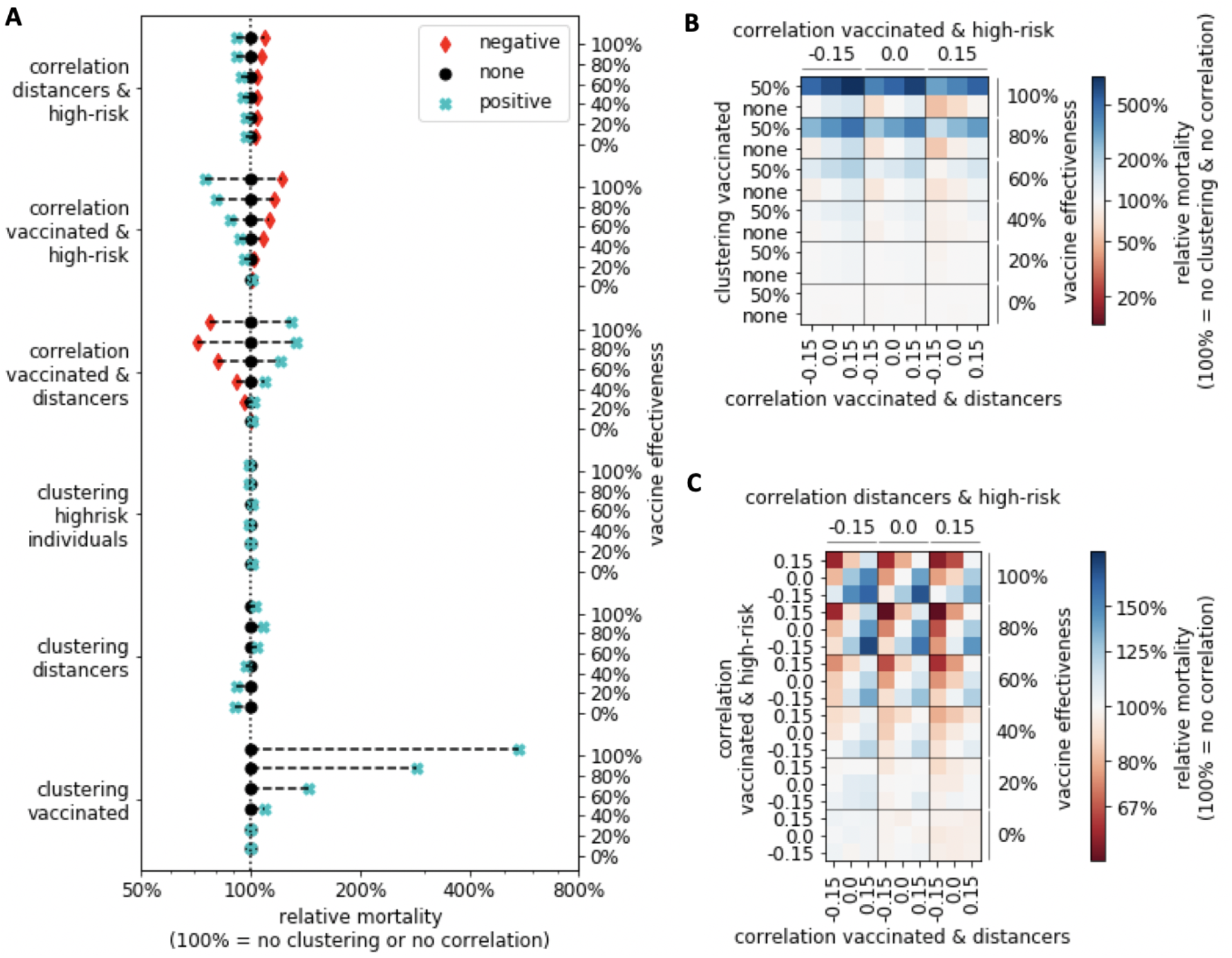
Relative mortality in the COVID-19 model compared to the homogeneous case of no clustering and no correlation. (A) For each line, the vaccine effectiveness and one clustering or correlation variable is fixed at a negative (− 0.15 correlation; red diamond), positive (0.15 correlation or 50% relative clustering; cyan cross) or zero value (black circle), and average mortality is calculated across all other values and compared to the case of no clustering or correlation (dotted line; relative mortality = 100%). (B-C) Relative mortality is shown when three variables and the vaccine effectiveness are fixed. In (B) the three most influential variables from (A) are fixed, while in (C) the three correlations are fixed. Blue (red) indicates higher (lower) mortality than in the homogeneous case of no clustering and no correlation.

Empirical studies in multiple countries indicate that older people have fewer daily physical interactions than younger people [21, 22]. As older people make up a large proportion of the group of high-risk individuals it makes sense to assume a lower contact rate, or alternatively a higher contact reduction rate for high-risk individuals compared to low-risk individuals. In addition to this inherent demographic difference, during the current pandemic contact reduction may be enhanced by additional social-distancing choices related to the higher perceived risk a COVID-19 infection presents for this vulnerable population. Without high-risk contact reduction, deaths can be averted by prioritizing the vaccination of high-risk individuals (Figure 5). To study the level of additional high-risk contact reduction at which the optimal vaccination strategy changes, we compared COVID-19-related mortality under three scenarios: high-risk individuals are more, equally and less likely to vaccinate than low-risk individuals. As expected, a reduction of contacts by high-risk individuals led to fewer deaths under all scenarios (Figure 6A). The rate at which deaths decreased varied significantly, however: if high-risk individuals strongly reduced their contacts, vaccination of proportionately more low-risk individuals led to fewer deaths (Figure 6B).

**Figure 6:**
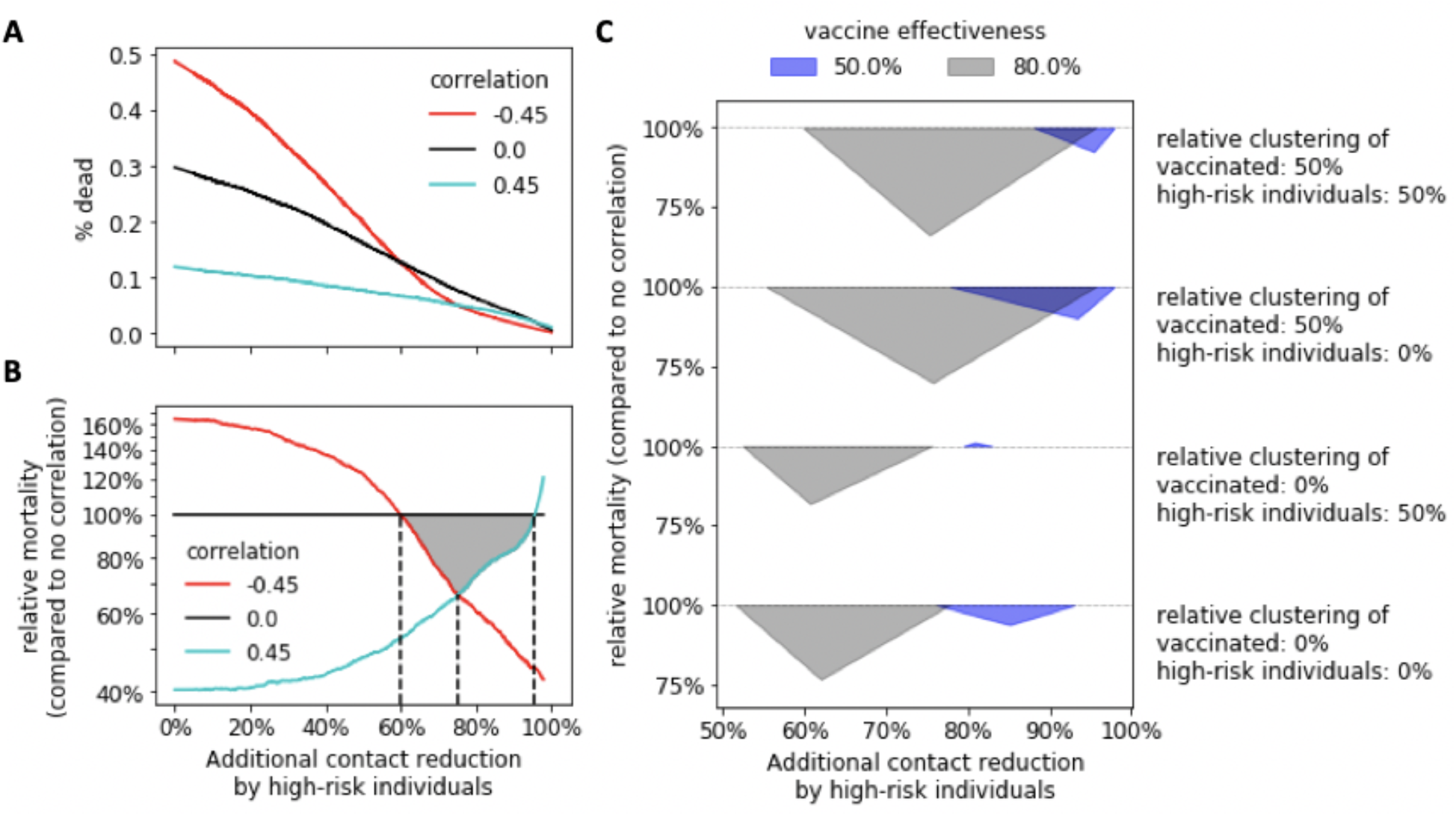
Level of contact reduction by high-risk individuals influences vaccination priorities. (A) The average mortality at a given additional contact reduction by high-risk individuals is shown for three different scenarios: negative (− 0.45; red), zero (black) and positive (0.45; blue) correlation between vaccinated and high-risk individuals. (B) Relative mortality compared to the case of no correlation (black line in A), at 50% relative clustering of both high-risk individuals and individuals who vaccinate. Black dashed lines and a gray triangle highlight the three intersection points of the three curves. (A-B) relative clustering of those who vaccinate and of high-risk individuals: 50%, vaccine effectiveness: 80%. (C) The location of the intersection points from (B) is shown for all combinations of relative clustering of those who vaccinate (0% vs 50%) and of high-risk individuals (0% and 50%), as well as two levels of vaccine effectiveness: 50% (blue) and 80% (gray). Figure S3 contains the full curves for all eight considered combinations.

We expected to find a single value for the additional contact reduction of high-risk individuals at which the correlation between who vaccinated and risk group did not matter. Instead, we found a range of values for which both negative and positive correlations yielded fewer deaths than no correlation. That is, a homogeneous vaccination strategy across risk groups (correlation = 0) was never optimal. For instance, in a situation with 80% vaccine effectiveness and 50% relative clustering of those who vaccinate and of high-risk individuals, we found that if high-risk individuals have 75% fewer interactions than low-risk individuals, both increased vaccination of low- or of high-risk individuals (correlation = 0.45) led to 34% fewer deaths than a homogeneous vaccination strategy (Figure 6B). As with other model results, the level of clustering of vaccinated individuals mattered, as the switch-point for optimal vaccination strategy was lower at 0% clustering of vaccinated versus 50% relative clustering. Furthermore, the switch-point was higher at lower vaccine effectiveness (Figure 6C). Interestingly, in the case of 0% relative clustering of vaccinated, 50% clustering of high-risk individuals, and a weak vaccine (effectiveness = 50%), if high-risk individuals reduced their contacts by 81% more than low-risk individuals, then the distribution of the vaccine across the risk groups did not affect mortality. This was the only parameter setting of those we considered that matched our original expectation.

## Discussion

Contact between similar people occurs at a higher rate than among dissimilar people, a phenomenon known as homophily [23]. While there is plenty of evidence that people with a negative view of vaccines cluster [24, 4, 25], we still lack empirical data on the degree of homophily regarding social distancing in response to the COVID-19 pandemic. There are, however, studies examining the average number of daily contacts stratified by age that show increased activity levels between people of similar age, which, given the strong correlation between COVID-19 risk status and age, suggests the presence of homophily regarding risk status. The correlation between policy views and partisan identification has been increasing [10], with more people now on the left or the right and fewer holding a mix of positions [11]. Recent polls by the Gallup agency indicate that an individual?s willingness to receive a COVID-19 vaccine [15] and to practice social distancing (wear a mask [12], eat out [13], fly [14], etc.) differs substantially by age as well as political party affiliation. Older people (age 65+), who are more likely at high risk, practice more social distancing and are slightly more willing to get vaccinated against COVID-19. Democrats are much more likely to receive the vaccine and to practice social distancing, which suggests a positive correlation between these two attributes considered in our study. We do not have accurate estimates for the degree of homophily or the correlation among different attitudes related to the spread of an infectious disease generally, or COVID-19 specifically. We therefore studied the spread of a disease across a social network under different possible scenarios for homophily and belief correlation.

We found that the presence of homophily in opinions regarding whether to vaccinate or not, and whether to practice social distancing or not, as well as a large overlap between those who distance and those who vaccinate, can dramatically increase the probability of a disease outbreak, especially in the presence of a highly effective vaccine. Accordingly, any results obtained using classical differential equation models, which inherently assume homogeneous mixing and account for neither homophily nor correlation in opinion patterns, likely present lower bounds on the expected severity of an outbreak. Furthermore, if opinions are positively correlated and if real interaction networks exhibit even a modest degree of homophily, as the Gallup surveys suggest they do, our current social context corresponds to a scenario with a substantially worse disease burden than a homogeneous scenario (Figure 3,5, S2).

Our study produced several results relevant to policy makers. First, whether vaccination of low- or high-risk individuals should be prioritized depends on the relative contact rate of low-versus high-risk individuals. If high-risk individuals have substantially fewer contacts (61 − 95% depending on the considered scenario; Figure 6C), prioritizing the vaccination of low-risk individuals reduces overall mortality compared to homogeneous vaccination or vaccination of high-risk individuals. The reason for this is likely that increased vaccination of low-risk individuals, who are more socially active, can prevent outbreaks and reduce mortality of the vulnerable, less-vaccinated population. An empirical study in eight European countries revealed that prior to the pandemic older adults (65+) already had on average 41.8% fewer contacts than those age 65 and under [21] (Table S2). Together with the increased perceived risk COVID-19 presents to the high-risk group, these individuals might indeed practice enough social distancing so that prioritizing the vaccination of low-risk individuals is optimal. Policy makers therefore might consider a heterogeneous approach, in which vaccination of high-risk individuals is prioritized in communities where social distancing among the high-risk population is less prevalent. Our findings highlight the importance of accurate estimates of the contact and risk reduction practiced by individuals of different risk groups. Second, the optimal vaccination strategy in our model was affected by the homophily among those who vaccinate. For a given vaccine effectiveness, the level of additional contact reduction at which the optimal vaccine strategy switches from prioritizing high-risk to prioritizing low-risk individuals is higher in the presence of homophily (Figure 6C).

Third, if vaccinated individuals engage in risk compensation, i.e., increase their activity levels, the level of effectiveness a vaccine needs to possess so that it does not lead to a worse disease burden increases in the presence of homophily and correlation, but only marginally. Finally, we note that almost all effects (both positive and negative) described in this study became stronger as the effectiveness of the vaccine increased. This may be partially due to the fact that situations with a vaccine effectiveness of 100% are on average closest to the herd immunity threshold of *R*_0_ = 1 (Figure S1, S2), as previous studies suggest the effect of homphily on outbreak probability is strongest when vaccination coverage is close to this threshold [5, 26]. Nevertheless, our results are counter-intuitive in this respect: While a more effective vaccine is certainly better in general, relative differences in outcomes and any negative effects due to the presence of homophily and/or correlation will be larger with a more effective vaccine.

We considered binary attributes and only second-order interactions among the attributes for two reasons: First, statistical methods for generating correlated Bernoulli random variables are well-established; and second, higher-order correlations lack an intuitive interpretation. Extending the model to include more binary attributes or higher-order interactions is straightforward using the algorithm we developed here. More complex scenarios could incorporate multinomial random variables with more than just two possible discrete values, random variables from other discrete distributions, or continuous random variables with appropriate supports. Simulating random variables from a joint distribution with a specified covariance structure can be technically challenging for many distributions, and research on this topic is ongoing [27, 28, 29]. The method we used to apply correlated random variables to nodes in a network to achieve appropriate clustering is readily extendable to multinomial random variables. The use of continuous random variables, however, would probably require a completely different algorithmic approach and a new measure of homophily. Further work in this area would expand the types of node attributes that could be modeled, and the types of correlations and clustering that could be studied.

There are theoretical limitations to the combinations of probabilities and correlations that can be generated (Figure S4). While the expected correlation is always 0, the range of compatible correlation values for given probabilities is not symmetric around 0 (e.g., two attributes, each with a high probability, can be strongly positively but not strongly negatively correlated). These limitations necessarily influenced the choices of probabilities and correlations we used in this study. In all analyses, we chose correlation values of equal magnitude so results could be compared in the positive and negative direction. Further, we compared correlations of magnitude 0.45 (0.15) when looking at two (three) binary attributes to ensure we investigate interaction networks where there are at least some individuals with each of the possible 4 (8) combinations of attributes. This means we studied only the effect of moderate and weak correlations. Stronger correlations will likely lead to stronger effects but exhibit the same directionality.

We modeled vaccine effectiveness using an all-or-nothing approach: either the vaccine provides full protection or it has no effect. A “leaky” vaccine that reduces the infection and/or the transmission probability for all vaccinated people by a certain percentage represents an alternative approach, however the model predictions may be insensitive to how vaccine effectiveness is implemented [30]. Modeling vaccine factors such as age-varying effectiveness requires the specification of further parameters, therefore to enhance the interpretability of our model we chose not to incorporate this additional level of complexity.

## Conclusion

People interact more frequently with people of similar age and with similar attitudes or beliefs, a phenomenon known as homophily. Surveys found that individuals at high-risk for severe COVID-19 infection are more likely to practice social distancing and get vaccinated once a vaccine is available, and positive attitudes toward social distancing and vaccination seem positively correlated. The current social context in which the COVID-19 pandemic is playing out is therefore characterized by homophily and correlations among beliefs and circumstances. We developed a novel technique for generating interaction networks with several binary attributes with a defined correlation structure and defined degrees of homophily. This technique is readily extendable to multinomial random variables, and can provide a foundation for more complex studies involving attributes with more flexible probability distributions.

We studied the spread of a generic infectious disease as well as COVID-19 in interaction networks with various levels of homophily and correlations among beliefs and circumstances. The current social context corresponds to a scenario with a substantially worse disease burden than a scenario without homophily and correlations. Differential equation models, which by nature assume homogeneous mixing of the population, likely underestimate the real disease burden. Several of our study results may be relevant to policy makers, as we showed that the optimal distribution strategy of a limited vaccine depends on the relative average contact rate of low-versus high-risk individuals, as well as the level of homophily between those who agree to be vaccinated.

## Methods

### Model summary

The modeling process consists of the following steps, described in detail in the following sections and depicted in Figure 1.

1. Generate a physical interaction network of *N* nodes.
2. For each node, draw *d* random binary attributes with a defined expectation and defined correlation structure,
3. Swap the values of an individual attribute between two nodes in order to increase the relative clustering of that attribute across the network. Stop when for each attribute a predetermined level of clustering has been achieved.
4. Vaccinate individuals willing to receive the vaccine and remove successfully vaccinated individuals from the pool of susceptibles.
5. Seed the network with a randomly infected individual.
6. Simulate the stochastic disease dynamics: transmission events, recovery, etc.
7. Record the initial reproductive number, if an outbreak occurred, and, in the COVID-19 model, the number of deaths.

### Physical interaction network

We studied the spread of a generic infectious disease (Figures 2-4) as well as COVID-19 (Figures 5-6) throughout a physical interaction network of *N* = 1000 individuals, modeled as a Watts-Strogatz small-world network [16]. In these networks, each vertex represents an individual and each edge represents a possible contact between two individuals. The average degree of each vertex was 14: the average number of contacts per day per individual found in a seminal multinational study (Table S2) [21]. We randomly rewired each edge with a probability of 5%, which is known to yield a high local clustering coefficient and low average path length, characteristic of small-world networks [16, 31].

### Modeling beliefs and circumstances

In the generic infectious disease model, we considered two binary attributes: attitude toward vaccination (1 = individual vaccinates, 0 = individual does not vaccinate), and attitude toward social distancing (1 = individual engages in risk reduction such as reduced interactions, mask wearing, increased hand washing, etc., 0 = individual does not engage in risk reduction). In the three-attribute COVID-19 model, we added risk status as a third variable (1 = individual is at high risk for COVID-19 due to age (65 and older) or known comorbidities, 0 = individual is not in the COVID-19 high-risk group).

### Generating binary attributes with a defined correlation structure

Exact forms for the distributions of correlated bivariate (*d* = 2) and trivarate (*d* = 3) Bernoulli random variables have been developed in the literature [32], and we relied on those results. We specified expectations *p*_*i*_,*i* = 1,…, *d*, and the covariance matrix **∑** and calculated a multivariate Bernoulli probability distribution of *d* correlated Bernoulli random variables. In the simplest case for *d* =2 (two binary attributes), if 

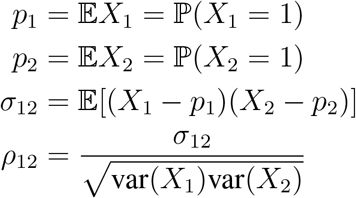

where *p*_*i*_ ∈ [0, 1], *ρ*_12_ ∈ [− 1, 1], and var(*X*_*i*_) = *p*_*i*_(1 − *p*_*i*_), the bivariate joint probability distribution can be represented as: 

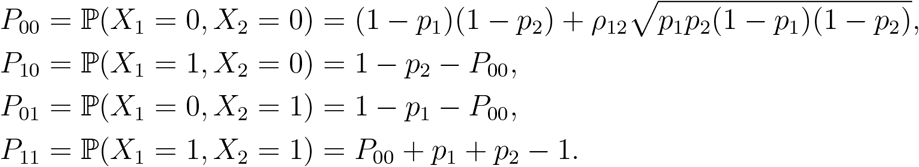

For a third random variable, *X*_3_, with random variables *X*_1_ and *X*_2_ as above: 

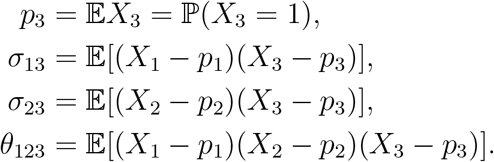

The multivariate distribution in three dimensions is given by 

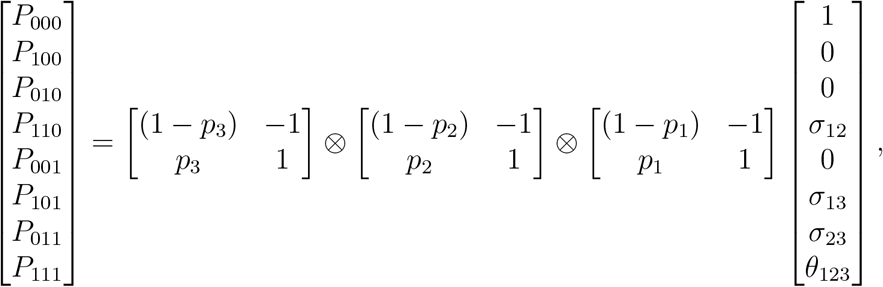

where ⊗ indicates the Kronecker product of two matrices. For all simulations in this study, we considered *θ*_123_ = 0. For *d* > 3 correlated binary attributes (not considered here), more computationally tractable algorithms have been developed [33, 34, 35], although the number of parameters required to fully specify the distribution is 2^*d*^ −1, and therefore grows exponentially.

One difficulty associated with generating correlated binary random variables has to do with the compatibility of the expectation vector and the covariance matrix. If **p** = [*p*_1_, *p*_2_,…*p*_*d*_] is a vector of expectations for d Bernoulli random variables, and **∑** is a covariance matrix, not all combinations of **p** and **∑** are compatible. For *d* = 2, an example of compatible correlation values which will result in a positive definite covariance matrix for fixed **p** is shown in Figure S4. Explicit bounds on the correlations for the case of *d* = 2 and *d* = 3 have been derived [36]. In general, as d increases, the probability that randomly chosen **p** and **∑** are compatible decreases quickly. For Figures 3, 4, 6 (two binary attributes), where we considered a fixed proportion of individuals who get vaccinated (*p*_1_ = 2/3) and who practice social distancing (*p*_2_ = 2/3), the compatible range was [− 0.5, 1] (Figure S4) and we considered correlation values − 0.45, 0, and 0.45. In Figure 4 (three binary attributes), the probability an individual gets vaccinated is *p*_1_ = 2/3, the probability an individual practices social distancing is *p*_2_ = 2/3, and the probability an individual is high-risk is *p*_3_ = 1/3. Here, the range of compatible correlation values was smaller, and we compared all combinations of second-order correlations of − 0.15, 0, and 0.15.

### Quantifying the relative clustering of binary attribute assignments in an interaction network

Given an assignment of a binary attribute (opinion or circumstance) *X* ∼ Bernoulli(*p*) to all *N* vertices (individuals) in a graph (interaction network), we counted the proportion of edges (interactions) between two vertices with the same attribute value (shared opinion or circumstance). Let this proportion be denoted by *ϕ* ∈ [0, 1]. Clearly, 𝔼 [*ϕ*] = *p*^2^ + (1 − *p*)^2^. Homophily is characterized by more interactions between individuals with shared attribute values than expected by random chance. We quantified the degree of homophily using the relative clustering of the binary attribute assignment, 

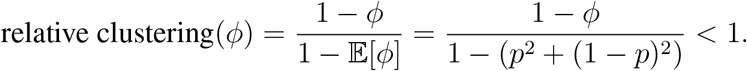

For example, if we assume that *p* = 2/3 of all *N* individuals get vaccinated, under a random expectation 𝔼 [*ϕ*] = 5/9 of all interactions will involve two individuals with the same vaccination status. Therefore, *ϕ* = 7/9 corresponds to 50% relative clustering.

The relative clustering value can be negative. However in this study we only considered non-negative levels of clustering since ‘heterophily’, the attraction by people with differing attribute values and/or the repulsion by people with same opinions or circumstances, seems unrealistic for the three binary attributes whose clustering effect we investigate here: who vaccinates, who practices social distancing and who is a high-risk individual. Furthermore, in a fully connected interaction network the relative clustering is always strictly less than 1 unless all individuals have the same opinion. However, in the limit as the network size N approaches infinity while the average degree remains fixed, the relative clustering can be chosen to be arbitrarily close to 1.

### Generating opinion patterns with a defined level of clustering and a defined correlation structure

We followed a two-step procedure to obtain an assignment of d binary attributes *X*_*i*_ ∼ Bernoulli(*p*_*i*_), i = 1,…, *d* to all *N* vertices in an interaction network with a predefined correlation structure between the attributes, as well as a predefined homophily (measured using relative clustering) for each attribute (Figure 1B-D).

1. To each vertex, we assigned a d-dimensional attribute vector with a defined correlation structure, by drawing from an appropriate multivariate Bernoulli probability distribution as described above.
2. We randomly picked one of the attributes that still exhibited lower than desired relative clustering. Then, we picked two vertices whose attribute vectors differed in only this attribute, and swapped their attribute values. This ensured that the correlation structure remained unchanged. We repeated this process until we reached the desired level of relative clustering for each attribute.

To ensure convergence of the latter process towards higher values of relative clustering, we defined the dissimilarity index of a vertex with respect to an attribute, denoted *d*(*v, a*), as the proportion of neighbors of this vertex with a different attribute value. We then preferentially picked vertices with a high dissimilarity index to swap attribute values.

To speed up the convergence process, we used (*d*(*v, a*))^16^ rather than the simple dissimarilty index *d*(*v, a*) to choose which two vertices to swap attribute values. This modification prevented the algorithm from converging to relative clustering values lower than desired, i.e., it ensured that we quickly reached high levels of relative clustering such as 50%. It did however slightly modify the resulting patterns of binary attributes as it led to fewer vertices with very high dissimilarity indices and relatively more vertices with intermediate index values. However, lower-sample-size simulations where we compared results obtained with an exponent of 16 to exponents of 1 and 4 revealed no qualitative differences in our findings (Figure S5).

### Effect of attribute values: vaccination, social distancing and high-risk individuals

Once attribute values were assigned to each individual, each vaccinated person was removed from the pool of susceptibles with a probability corresponding to the considered vaccine effectiveness.

Further, each individual was assigned a base activity level *a* ∈ [0, 1] describing how likely that individual was to seek contact with any neighbor in the interaction network on any given day. Throughout the paper, we used 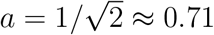, assuming that all individuals have fewer physical contacts that allow disease transmission than prior to a pandemic (due to e.g. mask wearing, work-from-home orders, etc.). The activity level of those individuals who practice social distancing was further reduced by *r*^distancing^ ∈ [0,1]. Note that this is a vertex-based attribute, implying that the probability of interaction between two people who practice social distancing was *r*^distancing^ · *r*^distancing^ lower than the probability of an interaction between two non-distancers. Also, note that while we primarily talk about social distancing throughout the paper, which can be easily understood in a network context, our abstract implementation of contacts does not require us to explicitly specify and separately model different types of risk mitigation. Rather, mask wearing, increased hand washing, social distancing, etc. all proportionately reduce the risk that a susceptible is infected through physical contact with an infectious individual, compared to a pre-pandemic level. The activity level of an individual should thus be interpreted as the combined effect of all risk mitigation efforts.

In Figure 4, we further considered the possibility of increased activity levels due to a potentially false belief of immunity following vaccination. We modeled this by introducing another vertex-based parameter *r*^increase^ ∈ [0, 1/*a* − 1], where the base activity level of vaccinated individuals is multiplied by 1+ *r*^increase^.

In the COVID-19 model, we added risk status as a third binary attribute, and considered clustering and correlation of this attribute in addition to vaccination and social distancing. High-risk individuals have a higher chance of a symptomatic, severely symptomatic, or deadly infection [17]. In Figure 6, we considered a continuum of scenarios where all high-risk individuals practice a certain increased level of distancing, modeled by multiplying the base activity level with *r*^distancing^ ∈ [0, 1] as before. In these analyses, the set of distancers coincides with the set of high-risk individuals and we did not consider differential distancing levels between individuals with the same risk status.

### Simulation of the disease spread

To simulate the spread of the generic infectious disease, we implemented a simple stochastic compartmental disease model and distinguished between susceptible (S), infected/infectious and removed/recovered (R) individuals. In the COVID-19 model, the infectious compartment was split into several compartments enabling a more accurate description of the course of COVID-19 progression: exposed/pre-symptomatic (E), asymptomatic (A), symptomatic (I), and severely symptomatic requiring hospitalization (H). To account for mortality, an additional removed compartment of individuals who have died from COVID-19 (D) was introduced [17]. In both models, time is discrete; one unit of time corresponds to one day. The simulation starts with a single seed case: a susceptible who becomes infected. In the generic infectious disease model, the seed case starts in the infected compartment, while in the COVID-19 model the seed case starts in the exposed compartment. The probability that any susceptible becomes the seed case is proportional to the activity level of that individual.

Each day, any susceptible can become infected through contact with an infectious neighbor on the interaction network. The probability of physical contact is the product of the activity levels of the susceptible and the infectious individual. If there is contact, then disease transmission occurs with transmission probability *β*. In the generic disease model, *β* = 10%. In the COVID-19 model, *β* varies over the course of the infection, is higher for symptomatic versus asymptomatic individuals and peaks at the onset of possible symptoms (details are described in [17]). Given the inherent uncertainty of COVID-19-related parameters, we sampled the transmission-related parameters from the same uniform probability distributions as in [17]: the peak transmission probability (provided contact occurs) for symptomatic individuals is *β*_*I*_ ∈ Unif([5%, 40%]) and for asymptomatic individuals *β*_*A*_ ∈ Unif([0%, /*β*_*I*_]).

In the generic infectious disease model, infectious individuals eventually recover from the disease, and the per-day probability of recovery is *γ*= 10%. In the COVID-19 model, infectious individuals start in the exposed compartment, while risk-group-dependent parameters and probabilities determine the transitions through the different infectious compartments. Infected individuals eventually recover or die. We used the same probability distributions and parameters governing these transitions as in the original model description [17]. Because of the short time frame of the simulations (weeks to months), we did not consider reinfections; recovered or dead individuals were removed from the simulation.

The more complex COVID-19 model includes additional parameters specific to SARS-CoV-2 and COVID-19, such as the proportion of asymptomatic infections and the infection fatality rate. We used the same values and sampled unknown parameters from the same uniform probability distributions as in the original model description [17].

### Outcome measures

In each simulation run of the generic infectious disease model, we recorded two outcome measures. First, an outbreak occurs if more than 1% of the population became infected (that is, at least 10 follow-up infectious occurred in a contact network of *N* = 1000 individuals). Second, we computed the initial basic reproductive number R_0_ as the total number of secondary infections caused by the seed case, i.e., th.0e initially infected individual. For this calculation, if on a given day a susceptible individual was “infected” by m > 1 people where one these people was the seed case, then we added 1/m to the initial basic reproductive number.

In the COVID-19 model, we recorded the total number of individuals who eventually died from the disease as an additional outcome measure. The outbreak probability and the initial basic reproductive number are unaffected by who is a high-risk individual. The severity of COVID-19 depends on risk status [20], and total mortality is the only outcome measure of the three we recorded that allows us to analyze the effect of homophily and correlation regarding risk status.

### Quantitative analysis

All model analyses were run entirely in Python 3.7. The contour plots in Figures 2,4 and S1 were generated by binning the data using a 20×20 equidistant grid, and subsequent smoothing using a 2-dimensional Savitzky-Golay filter [18]. To avoid over-smoothing, we chose a small window size of 3 and used only linear functions. Similarly, we used a one-dimensional Savitzky-Golay filter with window size 33,333 and linear functions to obtain a generalized moving average of the 333,333 data points generated for each correlation value in Figures 6,S3.

Each data point in Figures 3,5 and S2 represents an average value across all conducted simulation runs (10,000,000 in Figures 3 and S2,S5 and 1,000,000 in Figures 5 and S5), where those parameters shown on respective axes received a fixed value.

## Data Availability

The manuscript describes a computational study. No data was created.

## Notes

### Competing Interest Statement

The authors have declared no competing interest.

### Funding Statement

The authors received no specific funding for this work.

