## Supplementary Tables 1-2 and Supplementary Figures 1-5 for "Effect of clustering and correlation of belief systems on infectious disease outbreaks"

| relative clustering of vaccinated | relative clustering of distancers | Correlation | proportion vaccinated | proportion distancers | increase in activity levels by vaccinated | | | |
|---|---|---|---|---|---|---|---|---|
| | | | | | 10% | 20% | 30% | 40% |
| 0 | 0 | 0 | 0.5 | 0.5 | 14.8 | 25.3 | 32.1 | 42.4 |
| 0 | 0 | 0 | 0.5 | 0.65 | 16.8 | 23.5 | 35.6 | 40.2 |
| 0 | 0 | 0 | 0.5 | 0.8 | 10.7 | 25.3 | 32.8 | 40.6 |
| 0 | 0 | 0 | 0.65 | 0.5 | 20.1 | 28.3 | 37.2 | 41.3 |
| 0 | 0 | 0 | 0.65 | 0.65 | 15.3 | 27.1 | 34.3 | 40.9 |
| 0 | 0 | 0 | 0.65 | 0.8 | 14.1 | 24.2 | 33.1 | 40 |
| 0 | 0 | 0 | 0.8 | 0.5 | 19.6 | 28.4 | 33.9 | 39.2 |
| 0 | 0 | 0 | 0.8 | 0.65 | 14.9 | 26.2 | 33.2 | 37.8 |
| 0 | 0 | 0 | 0.8 | 0.8 | 14.4 | 25.9 | 34.8 | 39.4 |
| 0.5 | 0.5 | 0.45 | 0.5 | 0.5 | 20.8 | 32.1 | 43.8 | 45.1 |
| 0.5 | 0.5 | 0.45 | 0.5 | 0.65 | 24.3 | 32.3 | 39.6 | 45 |
| 0.5 | 0.5 | 0.45 | 0.5 | 0.8 | 15.7 | 28 | 36.3 | 46.7 |
| 0.5 | 0.5 | 0.45 | 0.65 | 0.5 | 14.7 | 28.5 | 32.6 | 39.1 |
| 0.5 | 0.5 | 0.45 | 0.65 | 0.65 | 16 | 29.9 | 36.4 | 42.7 |
| 0.5 | 0.5 | 0.45 | 0.65 | 0.8 | 17.8 | 27.7 | 36.9 | 43.7 |
| 0.5 | 0.5 | 0.45 | 0.8 | 0.5 | 16.9 | 28.7 | 34.3 | 41 |
| 0.5 | 0.5 | 0.45 | 0.8 | 0.65 | 20.8 | 30.5 | 35.6 | 41.8 |
| 0.5 | 0.5 | 0.45 | 0.8 | 0.8 | 17.1 | 25 | 34.9 | 41.7 |

Table S1: **Required effectiveness for a vaccine not to yield more outbreaks given an increased activity level by those who receive the vaccine.** For different proportions of those who vaccinate and those who distance ($50\%, 65\%, 80\%$) and two scenarios regarding clustering and correlation (none versus high clustering and correlation), $N = 200,000$ simulations each were conducted for each considered level of increased activity by those who vaccinated ($10\%, 20\%, 30\%, 40\%$) with randomly chosen vaccine effectiveness, $U([0\%, 100\%])$, in addition to $200,000$ simulations each without a vaccine. Using a one-dimensional Savitzky-Golay filter with window size $20,000$ and linear functions, we obtained smoothed plots of the outbreak probability against the vaccine effectiveness for each increased activity level by vaccinated, and inferred the respective vaccine effectiveness at which the outbreak probability under scenarios with a vaccine and increased activity levels by the vaccinated equalled the outbreak probability without a vaccine (green cell values).

| country | UN population data from year | average number of contacts per day by individuals < 65 years old | average number of contacts per day by individuals ≥ 65 years old | average contact reduction |
|---|---|---|---|---|
| Belgium | 2019 | 12.11 | 7.57 | 37.44% |
| Germany | 2018 | 8.05 | 5.97 | 25.81% |
| Finland | 2018 | 11.29 | 5.70 | 49.47% |
| Great Britain | 2018 | 11.57 | 7.93 | 31.47% |
| Italy | 2018 | 19.71 | 12.54 | 36.39% |
| Luxembourg | 2019 | 18.13 | 8.06 | 55.55% |
| Netherlands | 2019 | 15.34 | 8.05 | 47.50% |
| Poland | 2019 | 16.37 | 9.68 | 40.85% |
| unweighted average | | 14.07 | 8.19 | 41.80% |

Table S2: **Average daily contacts per country and age group.** Data from [21]. The most recently available census estimate from the United Nations Demographic Statistic Database was used for a weighted average of the contact rate across different age groups. The average contact reduction (last column) is calculated as one minus the ratio of average daily contacts by older people (fourth column) over the average daily contacts by younger people (third column).

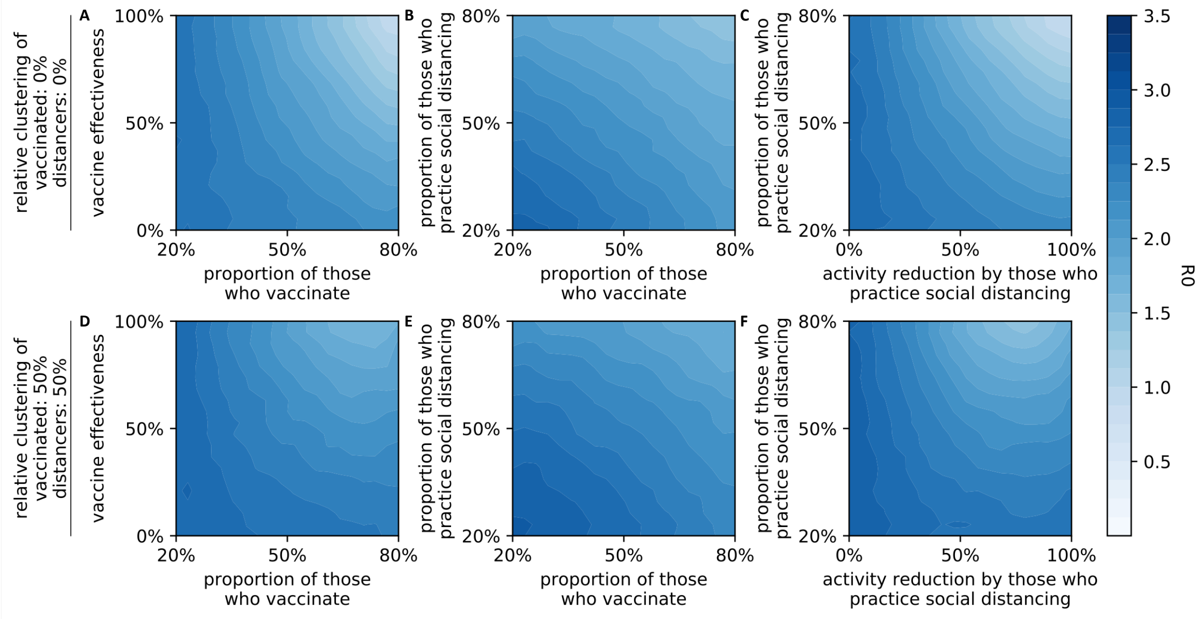


Figure S1: **Effect of vaccine and social distancing parameters on the basic reproductive number ($R_0$).** Contour plots were generated from 5,000,000 independent simulation runs with four vaccine and social distancing parameters chosen uniformly at random (axes show parameter ranges). Relative clustering levels of those who vaccinate and of those who practice distancing were fixed at (A-C) $0\%$ and (D-F) $50\%$. Data was binned and smoothed using a two-dimensional Savitzky-Golay filter [18] (details in Methods).

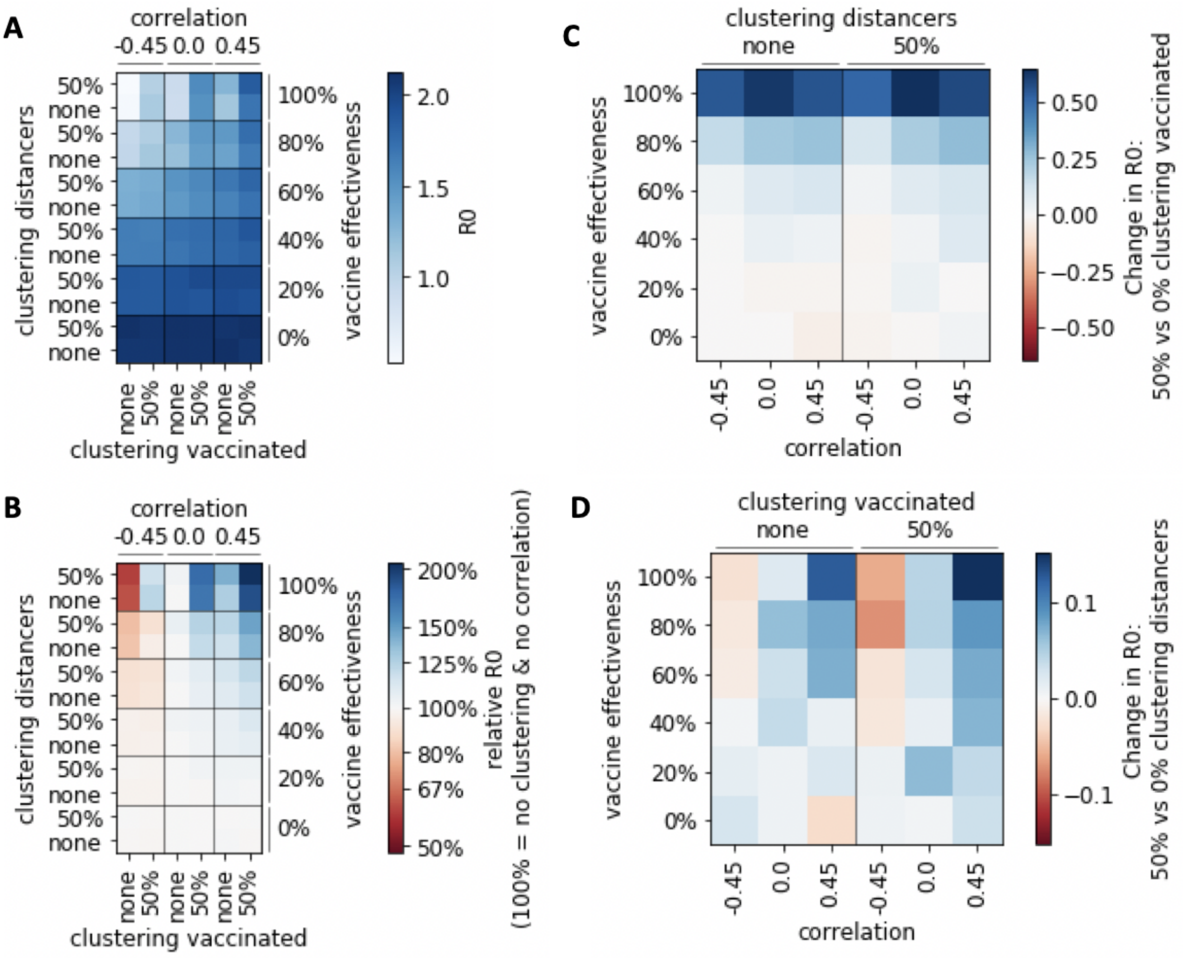


Figure S2: **Effect of clustering and correlation of opinions on the basic reproductive number.** (A-B) Basic reproductive number ($R_0$) under different scenarios regarding clustering and correlation of those who vaccinate and those who distance, and for different levels of vaccine effectiveness. (A) $R_0$, (B) relative change in $R_0$ compared to the homogeneous case of no clustering and no correlation. (C-D) Absolute change in $R_0$ when comparing physical interaction networks where (C) vaccinated, (D) distancers cluster versus networks without clustering of (C) vaccinated, (D) distancers.

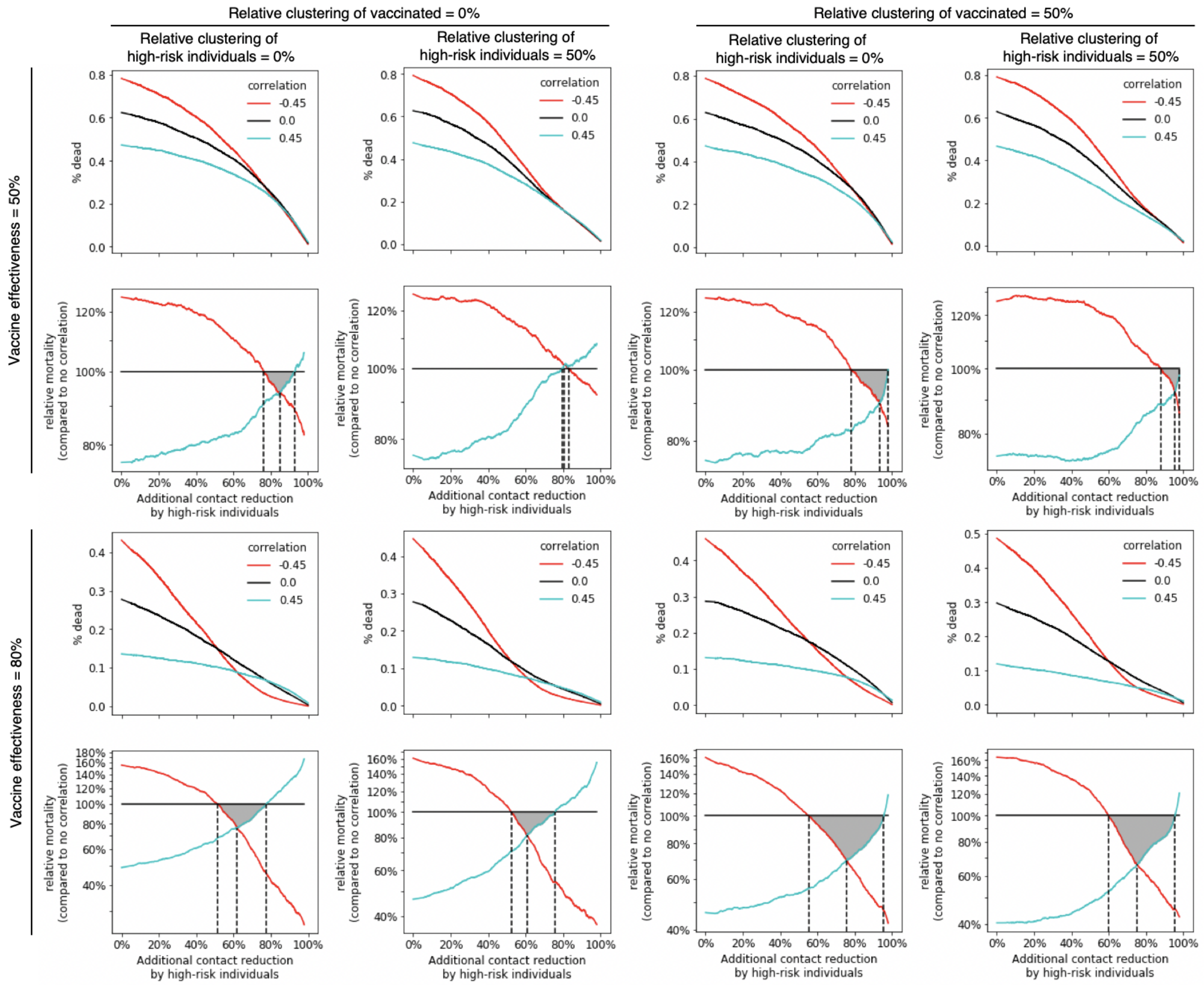


Figure S3: **Degree to which the level of contact reduction by high-risk individuals influences vaccination priorities under various scenarios.** The average absolute mortality (first and third row) at a given additional contact reduction by high-risk individuals is shown for three different scenarios: negative ($-0.45$; red), zero (black) and positive ($0.45$; blue) correlation between vaccinated and high-risk individuals. In addition, the relative mortality compared to the case of no correlation (black line) is shown (second and last row). Black dashed lines and a gray triangle highlight the three intersection points of the three curves. Different situations are considered: $50\%$ (first two rows) vs $80\%$ (last two rows) vaccine effectiveness, $0\%$ (first two columns) vs $50\%$ (last two columns) relative clustering of those who vaccinate, and $0\%$ (first and third column) vs $50\%$ (second and last column) relative clustering of high-risk individuals. For all eight scenarios, a direct comparison of the location of the gray region in between the intersection points is shown in Figure 6.

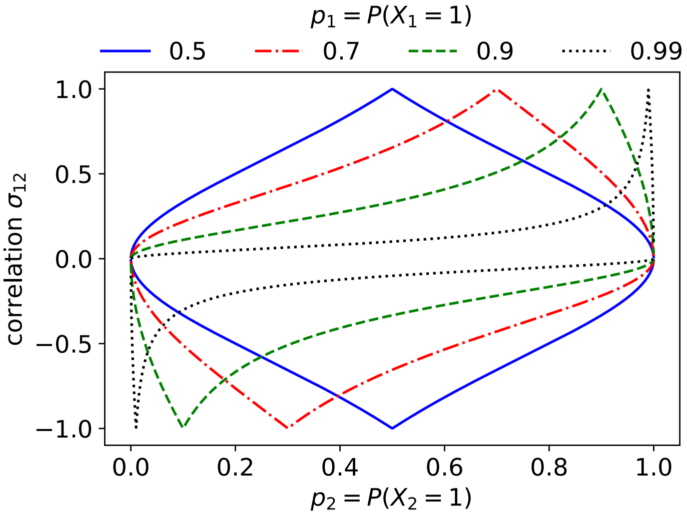


Figure S4: **Compatible choices for the expectation and correlation of two Bernoulli random variables.** The possible range of correlations between two Bernoulli random variables with expectations $p_1$ (colors) and $p_2$ (x-axis) is shown for four fixed choices of $p_1$.

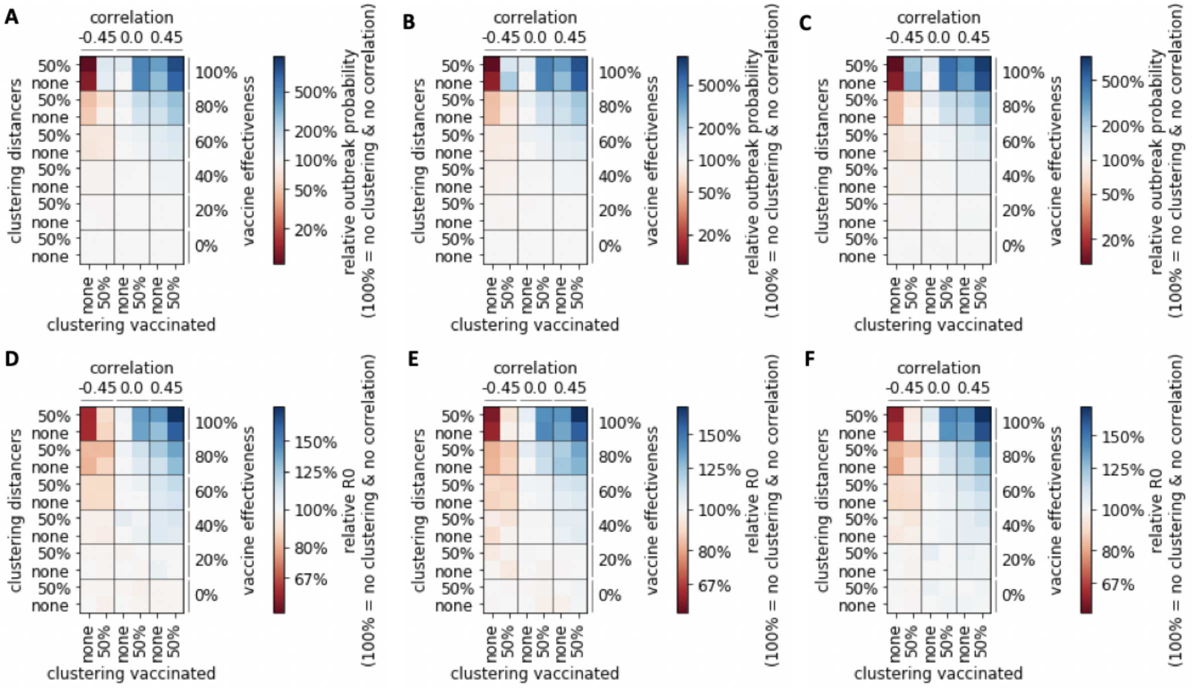


Figure S5: **Robustness of the results for various exponents in the clustering algorithm.** The relative change in (A-C) outbreak probability and (D-F) basic reproductive number $R_0$ compared to the homogeneous case of no clustering and no correlation is shown for different scenarios regarding clustering and correlation of those who vaccinate and those who distance, as well as for different levels of vaccine effectiveness. The exponent used in the clustering algorithm (see Methods) is $1$ in A and D, $4$ in B and E, and $16$ in C and F.
